# Model-based cost-effectiveness analysis of a multiplex lateral flow rapid diagnostic test for acute non-malarial febrile illness in rural South and Southeast Asian primary care

**DOI:** 10.1101/2023.10.28.23297707

**Authors:** Rusheng Chew, Chris Painter, Wirichada Pan-ngum, Nicholas Philip John Day, Yoel Lubell

## Abstract

**Background:** Multiplex lateral flow rapid diagnostic tests (LF-RDTs) may aid management of patients with acute non-malarial fever (NMFI) in rural South and Southeast Asia. We aimed to evaluate the cost-effectiveness in Cambodia and Bangladesh of a putative, as-yet-undeveloped LF-RDT capable of diagnosing enteric fever and dengue, as well as measuring C-reactive protein (CRP) to guide antibiotic prescription, in primary care patients with acute NMFI.

**Methods:** A country-specific decision tree model-based cost-effectiveness analysis was conducted from a health system plus limited societal perspective considering the cost of antimicrobial resistance. Parameters were based on data from a large observational study on the regional epidemiology of acute febrile illness, published studies, and procurement price lists. Costs were expressed in 2022 US dollars, and cost-effectiveness evaluated by comparing incremental cost-effectiveness ratios with conservative opportunity cost-based willingness-to-pay thresholds and the more widely used threshold of per capita GDP.

**Findings:** Compared to standard of care, LF-RDT-augmented clinical assessment was dominant in Cambodia, being more effective and cost-saving. The cost per DALY averted in Bangladesh was $482, slightly above the conservative opportunity cost-based willingness-to-pay threshold of $388 and considerably lower than the GDP-based threshold of $2,687. The intervention remained dominant in Cambodia and well below the GDP-based threshold in Bangladesh when antimicrobial resistance costs were disregarded.

**Interpretation:** These findings provide guidance for academic, industry, and policymaker stakeholders involved in acute NMFI diagnostics. While definitive conclusions cannot be made in the absence of established thresholds, our results suggest that cost-effectiveness is highly likely in some target settings and possible in others.

**Funding:** Wellcome Trust, UK Government, Royal Australasian College of Physicians, Rotary Foundation.

**RESEARCH IN CONTEXT:** *Evidence before this study:* The diagnosis and management of acute non-malarial febrile illness in rural South and Southeast Asian low- and middle-income countries remains challenging; multiplex lateral flow rapid diagnostic tests have been proposed as a possible solution. In view of the scant evidence on the epidemiology of acute non-malarial febrile illness in this region, we recently conducted an international Delphi survey to identify priority analytes for inclusion in such a putative test with capillary blood as the substrate. The main finding was that this test should be able to diagnose enteric fever and dengue, at a minimum; there was also support for including C-reactive protein as a means of differentiating viral from non-viral causes of NMFI to guide empirical antibiotic prescribing. However, we could not identify any economic evaluations of the cost-effectiveness of any such multiplex tests following a search of standard databases using the keywords ‘fever’, ‘febrile’, ‘multiplex’, ‘South Asia’ and ‘Southeast Asia’ on 2 March 2023.

*Added value of this study:* Bangladesh and Cambodia are lower-middle income countries in South and Southeast Asia, respectively, which are representative of the settings in which the test will be most useful. This country-specific assessment of the cost-effectiveness of such an urgently needed diagnostic tool provides vital information to guide decision-making by researcher, industry, and policymaker stakeholders on the development and deployment of such a test in terms of performance characteristics and pricing.

*Implications of all the available evidence:* The cost-effectiveness of this putative multiplex lateral flow rapid diagnostic test is dependent not only on its inherent performance and pricing, but on context-specific factors. Optimizing the specificity of component assays, as well as mechanisms to lower prices in markets where the test is not cost-effective, have the potential to ensure cost-effectiveness in as many target countries as possible.

## INTRODUCTION

In tropical low- and middle-income countries (LMICs), acute febrile illness is a common reason to seek healthcare.^1^ In such resource-limited settings, the majority of the population live rurally and usually present to public sector primary healthcare facilities in the first instance.^2^ Malaria was previously a very common aetiology of tropical acute febrile illness, but the success of malaria eradication programmes, especially in South and Southeast Asia,^3^ has exposed a large gap in the ability of primary healthcare workers to manage acutely febrile patients who test negative on malaria rapid diagnostic tests, most of whom will have other infections. Many of these non-malarial infections can be severe, but most are treatable yet often indistinguishable clinically.^4,5^ There is scant high-quality data on the regional burden of NMFI, but what little evidence there is suggests that it is large, with one surveillance study of Southeast Asian children showing an incidence density of 33.6 per 100 person-years.^6^

Several factors contribute to this lack of capacity. First, the many types of primary healthcare providers, which range from lay village health volunteers to clinics staffed by semi-skilled health workers, means that the workforce skill set relating to clinical diagnosis and management is variable but, in general, is limited.^2^ Second, there are a myriad causes of acute non-malarial febrile illness (NMFI), many of which present with non-specific symptoms.^1^ Third, while there are several rapid, point-of-care tests available, the vast majority are pathogen-based targeting one, or at most two, causes of NMFI, which is sub-optimal for effective patient management. The widespread uptake of these tests is hindered by well-described technical, biological, social, infrastructural, regulatory, and economic barriers.^7^ Where these are available, access is difficult as they are often found in secondary or higher-level facilities located a considerable distance from rural villages. Compounding this is the dearth of diagnostic tests suited for use in high temperature, high humidity rural primary care settings by low-skilled health workers.^8^

Multiplex multi-analyte rapid diagnostic tests, ideally analogous to those used for malaria, have been proposed as a possible solution to this problem.^9^ We recently conducted a modified Delphi survey to ascertain the analytes which should be included in such tests to aid management of acute NMFI in rural South and Southeast Asian settings, which would use capillary blood as the substrate and operate using lateral flow principles. The survey was performed given the paucity of robust regional epidemiological data relating not just to incidence, but also disease burden in terms of morbidity and mortality. The key finding was that these multiplex lateral flow rapid diagnostic tests (LF-RDTs) should, at the minimum, be able to diagnose acute enteric fever and dengue in patients of all ages, excluding neonates, with at least 75% sensitivity and 90% specificity. Additionally, C-reactive protein (CRP) was thought to be a useful analyte to include, given its ability to differentiate between bacterial and viral causes of acute febrile illness and, thus, promote antimicrobial stewardship by reducing inappropriate antibiotic prescriptions.^10^ This latter assertion is also supported by data from a study showing that a CRP level >40 mg/ml had a sensitivity of 74% for detecting bacterial infections in acutely febrile patients.^11^

While such a multiplex test has not yet been developed, assessment of its potential cost-effectiveness is essential to assist in guiding the research and policymaking processes that will underpin its successful development and implementation in the target settings. In this study, we aimed, therefore, to assess the cost-effectiveness in two tropical lower-middle income countries, one in Southeast Asia (Cambodia) and one in South Asia (Bangladesh), of a novel putative multiplex LF-RDT which measures CRP in addition to being able to diagnose acute enteric fever (e.g., by detecting typhoidal *Salmonella* antigens) and dengue (e.g., by detecting dengue NS1 antigen and dengue IgM) in patients with acute NMFI, in line with the results of the abovementioned Delphi survey.

## METHODS

### Setting

Cambodia is a lower-middle income country in mainland Southeast Asia with a population of 16·6 million, 75% of which live in rural areas, according to World Bank statistics from 2021.^12,13^ Bangladesh, in South Asia, is also a lower-middle income country but with a much larger population of 169·4 million and a rural population proportion of 61%.^14,15^ Cambodia and Bangladesh have young populations, with 29% and 26% aged <15 years, respectively.^16,17^ They also suffer from heavy burdens of enteric fever and dengue, prevalence estimates of which are shown in Table 1.

**Table 1.**
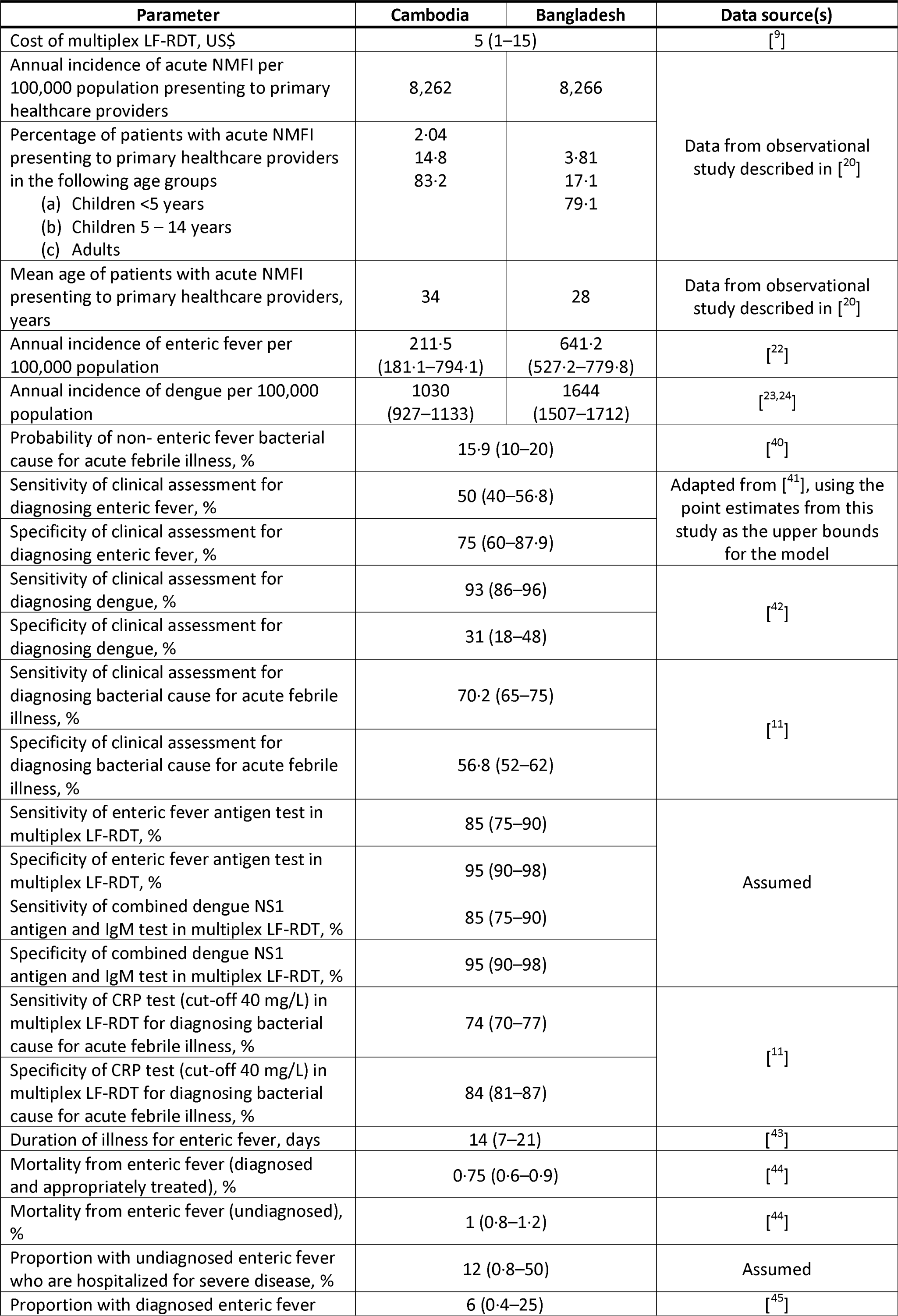
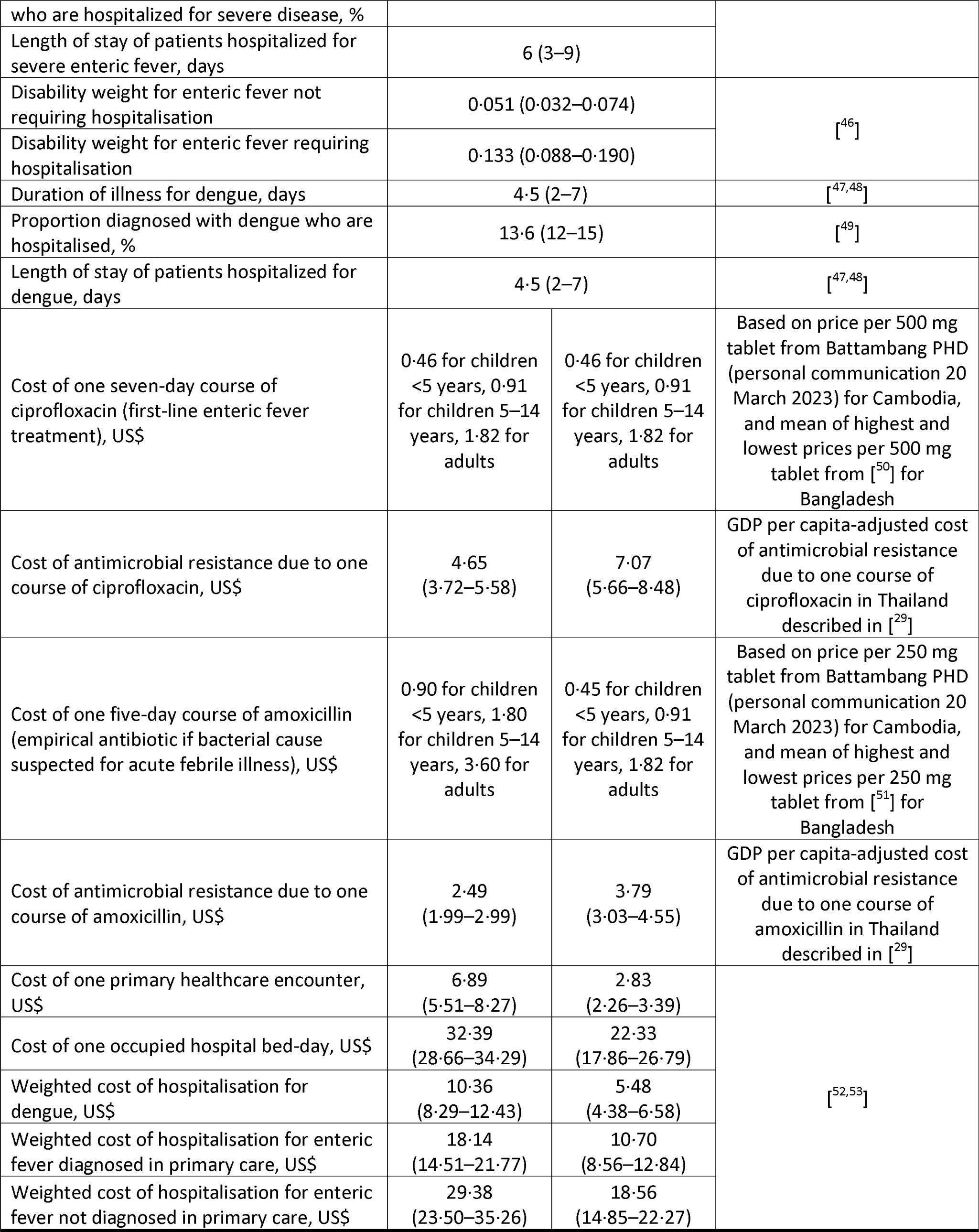
Decision tree model parameters for the main analysis. CRP, C-reactive protein; GDP, gross domestic product; PHD, Provincial Health Department; LF-RDT, lateral flow rapid diagnostic test. Costs were adjusted for inflation and expressed in 2022 US dollars.

In both these countries, the majority of rural primary healthcare is delivered by the government through a network of primary health centres (PHCs) and village health workers (VHWs),^18,19^ the latter mainly comprising lay people with minimal training. Like in many other lower-middle income countries, quality of care at PHCs and by VHWs is sub-optimal owing to low clinical skill levels e.g., inability to formulate syndromic diagnoses, and unavailability of diagnostics suited to these settings.

### Model design and participants

A model-based cost-effectiveness analysis was performed. This compared current management of acute NMFI, in which a diagnosis is made solely on clinical grounds without diagnostic aids, with a hypothetical counterfactual where management is augmented by a novel putative multiplex LF-RDT able to diagnose acute enteric fever and dengue using capillary blood as the test substrate, in addition to measuring CRP levels to aid in the differentiation of bacterial and viral causes of acute NMFI not attributable to enteric fever or dengue.

Models were based on 12 months’ worth of data collected between 21 March 2022 and 21 March 2023 for an ongoing large-scale observational study aiming to define the epidemiology of acute febrile illness in South and Southeast Asia. In this study, detailed clinical and epidemiological data, along with a more limited set of healthcare consumption and expenditure data were collected.^20^ Patients of all ages were consecutively recruited from among those presenting to rural PHCs and VHWs involved in the study, which were located in Battambang and Pailin provinces in Cambodia, and in Chittagong Division in Bangladesh. In brief, patients who presented acutely and who had an axillary temperature at presentation ≥37·5 or <35·5°C, or who had a history of fever in the 24 hours prior to presentation, and whose presentation was not due to accident or trauma and was not within three days of routine immunizations were eligible to participate.

For the purposes of this cost-effectiveness analysis, only participants older than 28 days residing in designated study villages (82 villages in Cambodia and 143 villages in Bangladesh) were included. In line with the use case for the multiplex LF-RDT, neonates were specifically excluded as fever in this age group generally requires assessment in hospital.^21^ The total populations of the Cambodian and Bangladeshi study villages were 65,567 and 115,962 persons, respectively. During the 12 month study period, 5,430 and 9,585 patients residing in study villages were recruited in Cambodia and Bangladesh, respectively. Dividing these by the total populations of the study villages yielded annual incidence estimates of patients with acute febrile illness who sought primary healthcare per 100,000 population of 8,262 in Cambodia and 8,266 in Bangladesh. All were tested with malaria rapid diagnostic tests but none returned a positive result in Cambodia and only 137 (1.4%) tested positive in Bangladesh. Aetiological data from this study are not yet available, hence published estimates for the incidence of enteric fever and dengue were used in the model.^22-24^ Age group-specific incidence estimates for the following age groups by country are shown in Table 1: children <5 years, children 5–14 years, and adults.

### Model structure

A decision tree model was constructed to compare the standard of care with multiplex LF-RDT-aided management for the abovementioned age groups. Like previous economic evaluations of other LF-RDTs in the South and Southeast Asian context,^25-28^ this model type was selected on the basis of the multiplex LF-RDT being similar to other well-established RDTs, such as those for malaria, in terms of ease of use and interpretation, short time to diagnosis, established acceptability to patients and health workers, and that the principal factor being influenced is clinical decision-making. The structure of the decision tree is shown in Figure 1, and a summary of the recommended interpretation and management for positive test components is shown in Supporting Information S1. Separate analyses were performed for Bangladesh and Cambodia. Model parameters were derived from a combination of data from the observational study as well as data from the published literature, and are shown in Table 1.

**Figure 1.**
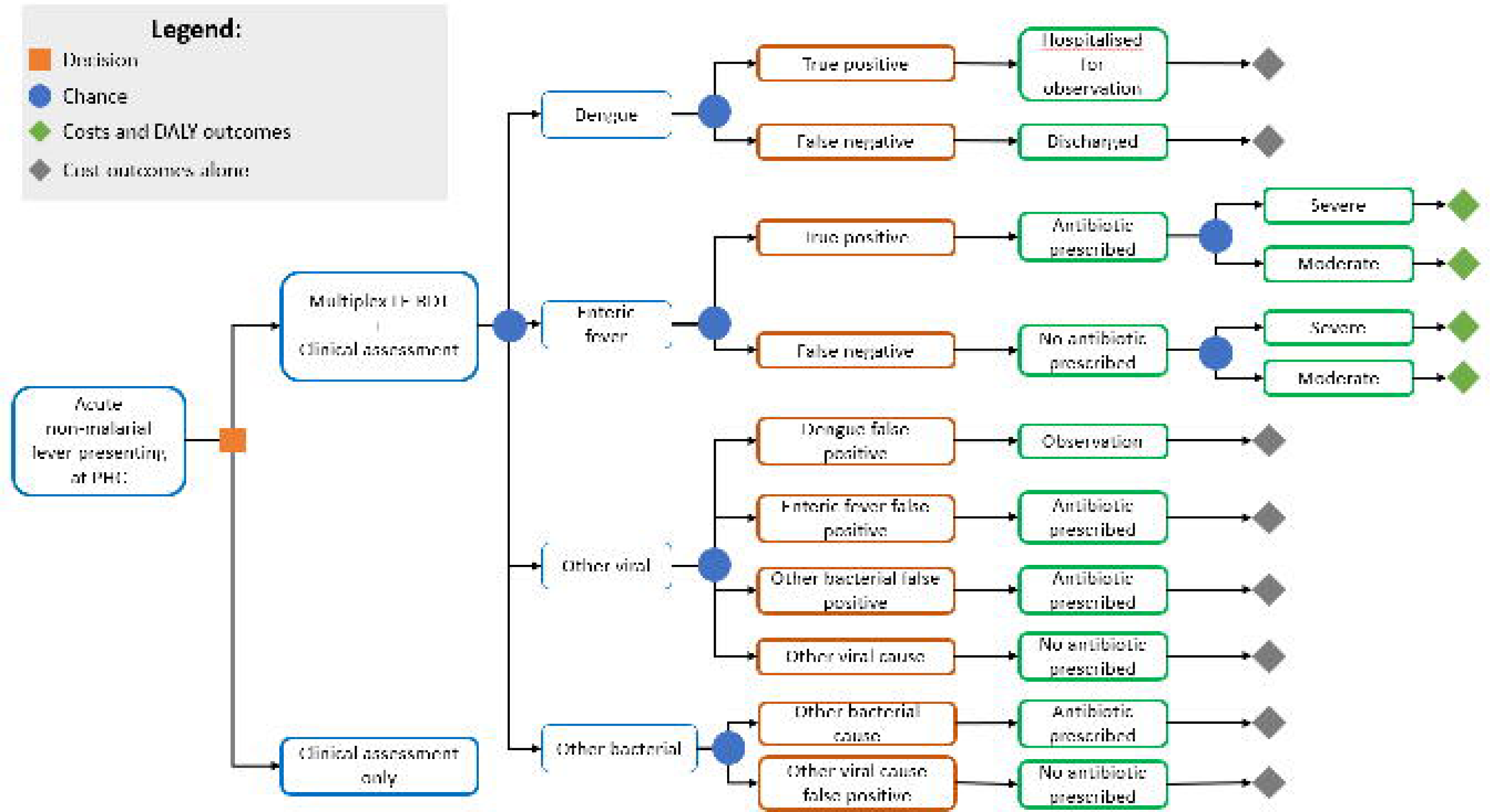
Decision tree model structure. The chance nodes arising from the ‘Clinical assessment only’ internal node are similar to those arising from the ‘Multiplex LF-RDT + Clinical assessment’ internal node. Parameters are shown in Table 1. LF-RDT, lateral flow rapid diagnostic test; PHC, primary health centre.

The outcomes of interest were disability-adjusted life-years (DALYs) averted for enteric fever, hospitalisations averted for dengue, total antibiotic prescriptions, inappropriate empirical antibiotic prescriptions averted, and total costs. A healthcare system plus limited societal perspective where costs of antibiotic resistance were incorporated was adopted. The latter cost was calculated using the three-component model constructed by Shrestha et al. with broad-spectrum penicillins and quinolones as the antibiotic classes of interest and adapted for the Cambodian and Bangladeshi contexts based on their gross domestic products (GDP) relative to that of Thailand (the context in which the estimates were derived); the three components are the correlation coefficients between human antibiotic consumption and subsequent resistance, the economic costs of antibiotic resistance for five sentinel pathogens, and consumption data for antibiotic classes driving resistance in these organisms.^29^ Acquisition costs of antibiotics were calculated for each age group to reflect the variation in antibiotic course prescriptions in each group.

Costs were adjusted for inflation to the 2022 cost year based on the relevant country Consumer Price Indices, and expressed in US dollars. The time horizon was one year, and no discounting or age-weighting was applied, in line with guidance established since the Global Burden of Disease 2010 report,^30^ and given the acute nature of the diseases of interest. Thus, although the modelled time period was one year, the DALYs accrued within this time period included the full years of life lost from the patient’s life expectancy. Cost-effectiveness was expressed as an incremental cost-effectiveness ratio (ICER), in terms of an incremental cost per DALY. Currently neither Bangladesh or Cambodia have explicit or accepted willingness-to-pay, or cost-effectiveness, thresholds. As such, the means of the opportunity cost-based thresholds for Bangladesh and Cambodia reported by Woods et al. were used and inflated to 2022 values ($357 for Cambodia and $388 for Bangladesh).^31^ For comparison, and given this uncertainty, the less conservative figure of the national per capita GDP was also used as a secondary willingness-to-pay threshold. This is in line with the lower bound for the willingness-to-pay threshold of one to three times per capita GDP recommended by the World Health Organization CHOosing Interventions that are Cost-Effective (WHO-CHOICE) programme, which does not take into account opportunity costs.^32^ The 2022 per capita GDP values for Cambodia and Bangladesh are $1,765 and $2,687, respectively.^33,34^

The analysis was performed using Excel (Microsoft, Washington, USA). The parameterized model and associated calculations for each country are shown in Supporting Information S1.

### Assumptions

Several assumptions were used in the model. First, the sensitivity and specificity of the multiplex LF-RDT for both enteric fever and dengue was assumed to be 85% (range 75– 90%) and 95% (range 90– 98%), respectively. These values were chosen bearing in mind the WHO-recommended performance characteristics for multiplex rapid diagnostic tests for acute febrile illness,^9^ the minimum performance metric requirements for a diagnostic assay to be clinically useful,^35^ and the constraints placed on sensitivity with a very small volume of substrate.^36^ This approach is well-established when performing economic evaluations of interventions.^37^ Second, ciprofloxacin was assumed to be the appropriate treatment for all cases of enteric fever, notwithstanding the increasing rates of drug resistance in the region.^38^ Third, full compliance by healthcare workers with multiplex LF-RDT-guided clinical management and by patients with their management plans was assumed. Fourth, amoxicillin was assumed to be the default antibiotic prescription for acute NMFI not caused by enteric fever and for which a bacterial cause is suspected. This assumption was made given that amoxicillin was the antibiotic most prescribed for undifferentiated fever in these settings based on data from the observational study,^20^ which is also in line with the limited formularies available in rural Cambodian and Bangladeshi primary care. Fifth, the cost of the multiplex LF-RDT was assumed to be $5, the optimal full cost to the payer recommended by the WHO for multiplex multi-analyte diagnostic tests for acute febrile illness.^9^ Cost containment is also aided by the operational similarity of the multiplex LF-RDT to malaria rapid diagnostic tests, and its rollout leveraging existing malaria control programme training and monitoring activities. Sixth, in the absence of detailed information on age-group specific disease incidence, the same country-wide incidence estimates were assumed to apply across all age groups. Seventh, it was assumed that undiagnosed (and, therefore, inappropriately treated) enteric fever cases were twice as likely to require hospitalisation than correctly diagnosed and treated cases, but in the absence of data it was conservatively assumed that correct diagnosis and treatment of enteric fever reduced mortality from 1% to 0·75%. Eighth, to avoid overcomplicating the model structure with all of the possible test result combinations from the multiplex LF-RDT, it was assumed that false positives could only arise from the ‘other viral infection’ group (e.g., no true dengue cases could produce false positive test results for enteric fever and vice versa). Lastly, the main use-case of the dengue component of the multiplex LF-RDT is to minimise inappropriate referrals to hospital. However, since it cannot predict development of severe dengue (which occurs around the seventh day of illness) and for which treatment is supportive only, we assumed that the intervention would have no mortality impact on dengue true positives and false negatives. Of note, while all patients diagnosed with dengue at the primary care level will be referred to hospital, only 13.6% will ultimately be admitted as some false-positives will be given alternative diagnoses and some true-positives will not be admitted.

### Sensitivity and scenario analyses

Deterministic (one-way) and probabilistic sensitivity analyses were performed. In the former, outcomes were assessed using the lower and upper estimates of each of the model parameters sequentially to assess the effect of uncertainty in individual parameters, displaying these as a tornado plot. For parameters without available confidence intervals and standard deviations to inform the upper and lower bounds, a default variation of 20% of the mean parameter value was assumed.

In the probabilistic sensitivity analysis, standard assumptions were made regarding the distribution of each parameter (β for individual probabilities and percentages, Dirichlet for multivariate probabilities and percentages, and γ for costs). Parameter values were randomly drawn from their respective distributions in a Monte Carlo simulation, with results from 1,000 model iterations displayed in the form of a scatter plot. Mortality from enteric fever was varied such that undiagnosed enteric fever would always have a value equal to or higher than that of diagnosed and appropriately treated enteric fever. A cost-effectiveness acceptability curve was also constructed to illustrate the how the estimated probability of the cost-effectiveness of the intervention changes at different willingness-to-pay thresholds.

Four scenario analyses were also performed. In the first, the percentage sensitivities and specificities of each multiplex LF-RDT component were varied in increments of 10% (range 60–90%) with the condition that they totalled 150%, which is the minimum requirement for a test to be clinically useful.^35^ This was first done singly while retaining the base case performance characteristics of the remaining two components, and then simultaneously for all three components. We ascertained average net monetary benefits per patient tested and threshold cost-effective prices for each of these test performance characteristic combinations to determine the most cost-effective potential combination, in the event the technical difficulties associated with producing such a multiplex assay precluded achievement of the performance characteristics described in the base case. The second assessed the impact future upskilling of primary health workers in clinical diagnosis would have on the cost-effectiveness of the multiplex LF-RDT. For this scenario, we assumed that health workers would be able to formulate syndromic diagnoses based on the criteria used in the observational study and that multiplex LF-RDTs would only be applied to acutely febrile patients with no localizing symptoms, details of which are shown in Table 2, in keeping with the syndromic presentation of the vast majority of enteric fever and dengue cases. In the third, the uncertainty in mortality benefit was evaluated through scenarios in which there was no benefit and a 50% reduction in mortality from enteric fever arising from use of the multiplex LF-RDT, respectively. Finally, the impact of the societal cost of antimicrobial resistance on cost-effectiveness was assessed by excluding this cost from the model.

**Table 2.** Data on incidence of non-malarial febrile illness with no localizing symptoms used to parameterize the model in a scenario where the intervention was only used in such patients. The remaining model parameters were those used in the base case analysis (see Table 1). NMFI, non-malarial febrile illness.

| Parameter | Cambodia | Bangladesh | Data source(s) |
| --- | --- | --- | --- |
| Annual incidence of acute NMFI with no localizing symptoms per 100,000 population presenting to primary healthcare providers | 5,259 | 955 | Data from observational study described in [<sup>20</sup>] |
| Percentage of annual incidence of acute NMFI with no localizing symptoms per 100,000 population presenting to primary healthcare providers in |  |  |  |
| (a) Children <5 years | 2·64 | 3·52 |  |
| (b) Children 5 – 14 years | 13·1 | 13·6 |  |
| (c) Adults | 84·3 | 82·8 |  |
| Mean age of patients with acute NMFI with no localizing symptoms presenting to primary healthcare providers, years | 34 | 28 |  |

This report was prepared in accordance with the Consolidated Health Economic Evaluation Reporting Standards (CHEERS) guideline.^39^ The completed CHEERS checklist can be found in Supporting Information S2.

## RESULTS

### Base case analysis

In Cambodia, compared to the current standard of care, augmentation of clinical assessment with the novel multiplex LF-RDT nearly tripled the proportion of correct diagnoses from 2,717/8,262 (33%) to 7,505/8,262 (91%) annually, leading to 497 (93%) fewer unnecessary hospitalisations for dengue-negatives. It also resulted in 523 (32%) fewer inappropriate antibiotic prescriptions, and 344 (13%) fewer antibiotic prescriptions overall. On a per-patient basis, hospitalisation costs were reduced from $7·93 to $2·20, costs attributable to antimicrobial resistance from $1·21 to $0·81, and total costs from $9·90 to $8·82. The number of DALYs averted per patient was 0·0012, resulting in the multiplex LF-RDT being dominant when compared with both lower and higher willingness-to-pay thresholds.

In contrast, the intervention more than doubled the number of correct diagnoses from 3,468/8,266 (42%) to 7,455/8,266 (90%) in Bangladesh, resulting in 548 (36%) fewer inappropriate antibiotic prescriptions, although the overall antibiotic prescription rate was little changed (29·3% with use of the multiplex LF-RDT vs. 30·6% without). There were 404 (93%) fewer unnecessary hospitalisations for dengue-negatives, helping to drive the average hospitalisation cost down from $4·70 to $2·11. The magnitude of antimicrobial resistance-related costs averted per patient was similar to that seen Cambodia at $0·40. In this setting, multiplex LF-RDT usage was associated with a slightly higher number of DALYs averted per patient at 0·0041 at an average total cost of $1·96, resulting in an ICER of $482 per DALY averted i.e., slightly higher than the lower willingness-to-pay threshold of $388 per DALY averted but well below the higher threshold of $2,687 per DALY averted. At the lower threshold, the threshold price for the multiplex LF-RDT to be cost-effective in Bangladesh was $4·62 per unit. Table 3 compares the key findings of the base-case analysis for Cambodia and Bangladesh.

**Table 3.** Key findings of the base-case analysis for Cambodia and Bangladesh. DALY, disability-adjusted life-year; LF-RDT, lateral flow rapid diagnostic test.

|  | Cambodia | Bangladesh |
| --- | --- | --- |
| Incremental cost per patient, \$ | -1.08 | 1.96 |
| DALYs averted per patient | 0.0012 | 0.0041 |
| Incremental cost per DALY averted, \$ | Cost-saving | 482 |
| Lower willingness-to-pay threshold (per DALY averted), \$ | 357 | 388 |
| Higher willingness-to-pay threshold (per DALY averted), \$ | 1,765 | 2,687 |
| Cost-effectiveness of multiplex LF-RDT | Dominant | Not cost-effective at lower willingness to pay threshold but cost-effective at higher threshold |

An important finding is that the increased ability to diagnose enteric fever correctly with the multiplex LF-RDT also reduced inappropriate empirical prescribing of ciprofloxacin considerably (by 70% in Cambodia and 47% in Bangladesh), moving the bulk of inappropriate antibiotic prescribing towards amoxicillin. This is beneficial because ciprofloxacin, with its much broader spectrum of activity compared to amoxicillin, is a bigger driver of antimicrobial resistance and, as such, is associated with a higher societal cost. Additionally, the intervention helps target ciprofloxacin prescriptions towards those in which they are most required i.e., patients with enteric fever.

### Sensitivity analyses

As can be seen in the results of the deterministic sensitivity analysis shown in Figure 2, Cambodia and Bangladesh share seven parameters impacting cost-effectiveness, but their relative importance differs greatly between the two countries. The vast majority of these factors do not relate to the multiplex LF-RDT. The effect of varying the unit price had the same impact on net monetary benefit in both countries, and was of primary importance in both countries respect to the cost-effectiveness thresholds. Differences in mortality between diagnosed and appropriately treated enteric fever and undiagnosed enteric fever had a greater impact than cost of healthcare provision in Bangladesh while the converse is true for Cambodia, reflecting the higher incidence of enteric fever in the former.

**Figure 2.**
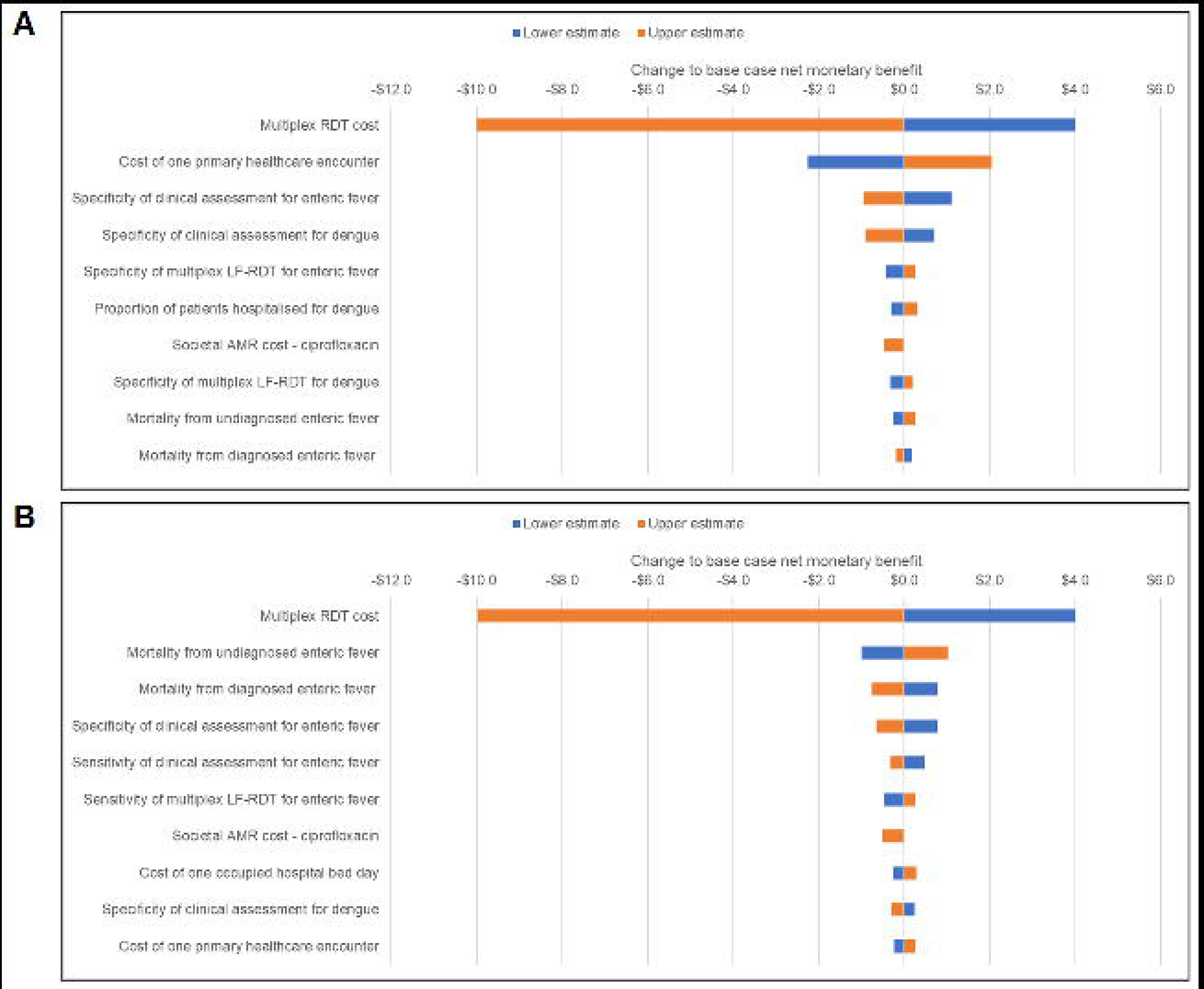
Tornado plots for (A) Cambodia and (B) Bangladesh showing the ten most important parameters affecting cost-effectiveness in each country. AMR, antimicrobial resistance; LF-RDT, lateral flow rapid diagnostic test.

The results of the probabilistic sensitivity analysis are shown in Figure 3. < majority of the incremental cost-effect simulation results, as well as the mean result, lie below the *x*-axis of the scatter plot, indicating that augmenting clinical assessment with the multiplex LF-RDT is likely to be cost-effective, with relatively little uncertainty regarding the number of DALYs averted [Figure 3A(i)]. This is reflected in the associated cost-effectiveness acceptability curve which shows that the intervention had a much higher probability of being cost-effective than standard of care at any willingness-to-pay threshold up to $10,000 per DALY averted [Figure 3A(ii)]. In other words, the probability that the data are consistent with the true cost-effectiveness ratio falling below this threshold is much higher for the intervention than standard of care. The opposite is true in Bangladesh, where the spread of incremental cost-effect pairs is more dispersed i.e., there is higher uncertainty [Figure 3B(i)]. The probability of the intervention being more cost-effective than standard of care in Bangladesh occurred at a willingness-to-pay threshold exceeding $500 per DALY averted, but the differences in probability of being cost-effective were considerable beyond this threshold [Figure 3B(ii)].

**Figure 3.**
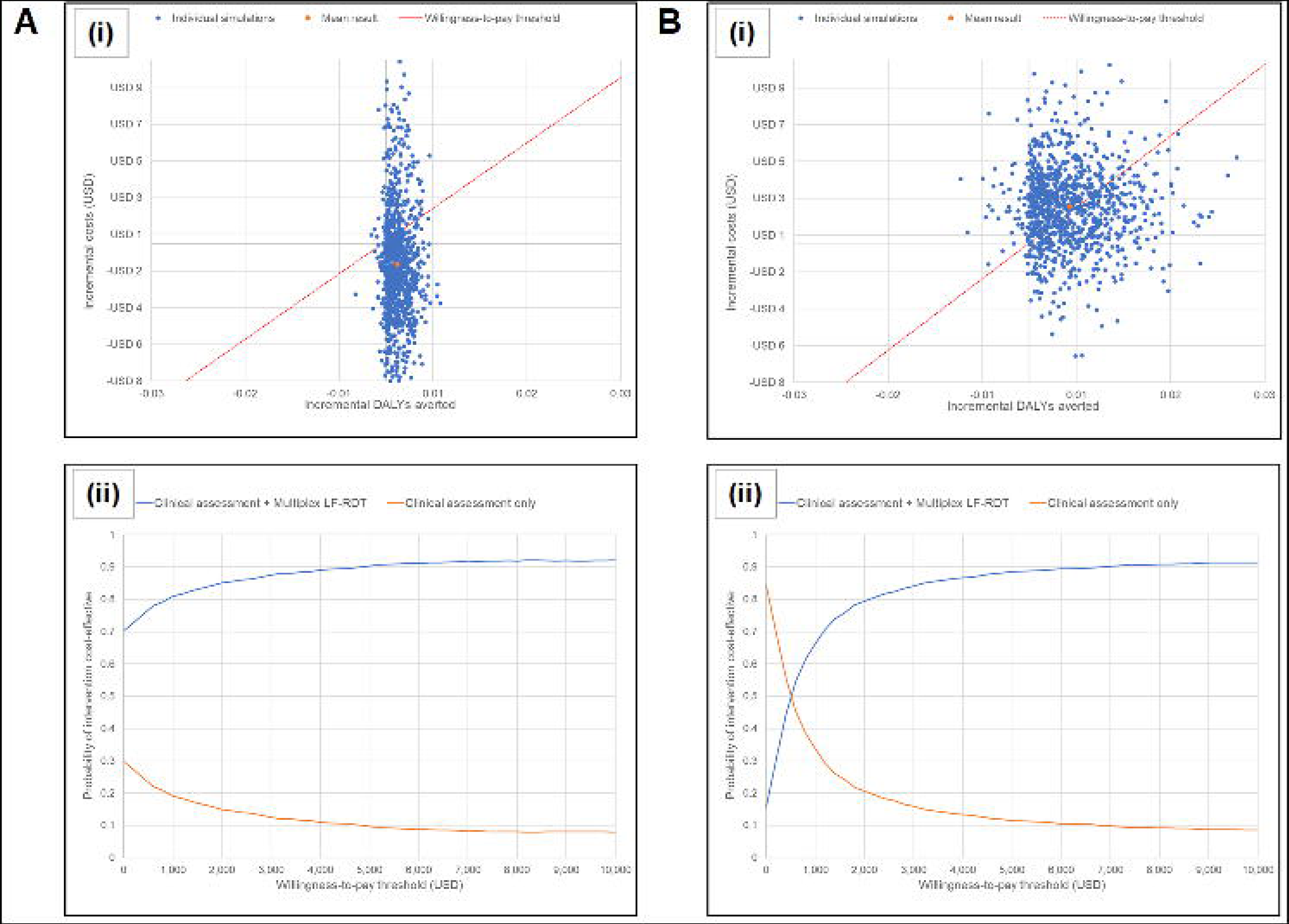
(i) Scatter plots of incremental cost-effect pairs for augmentation of clinical assessment with the multiplex LF-RDT obtained from 1,000 Monte Carlo model simulations randomly drawing parameter values from their respective distributions for (A) Cambodia and (B) Bangladesh. Standard assumptions were made regarding the distribution of each parameter (β for individual probabilities and percentages, Dirichlet for multivariate probabilities and percentages, and γ for costs). The conservative opportunity cost-based willingness-to-pay thresholds of $357 and $388 per DALY averted in Cambodia and Bangladesh, respectively were used. (ii) Cost-effectiveness acceptability curves for (A) Cambodia and (B) Bangladesh showing how the estimated probability of the cost-effectiveness of clinical assessment augmented by the multiplex LF-RDT vs. clinical assessment alone changes at different willingness-to-pay thresholds. DALY, disability-adjusted life-year; LF-RDT, lateral flow rapid diagnostic test; USD, US$ at 2022 values.

### Scenario analyses

When the sensitivity and specificity of the multiplex LF-RDT components were varied individually and together on the condition that they totalled 150%, increasing specificity of any component at the expense of sensitivity resulted in increasing net monetary benefit per patient tested, thus allowing a higher threshold cost-effective price for the multiplex LF-RDT (Table 4). This was particularly true for CRP, where a slight increase in specificity to 90% from 84% yielded average net monetary benefits greater than those seen in the base case analysis of $1·49 and -$0·38 in Cambodia and Bangladesh, respectively. Nevertheless, this was still insufficient to make the multiplex LF-RDT cost-effective in Bangladesh when the more conservative willingness-to-pay threshold was applied. Reducing the sensitivity of the dengue component from 85% to 60% and specificity from 95% to 90% also marginally increased cost-effectiveness in Bangladesh, although again this was insufficient to bring the ICER below the lower willingness-to-pay threshold. Even if all three components had the same sensitivity and specificity values of 60% and 90%, respectively, the multiplex LF-RDT would still be dominant in Cambodia and a little less cost-effective in Bangladesh when compared with the lower willingness-to-pay threshold.

**Table 4.** Effect of varying sensitivity and specificity for each multiplex LF-RDT component singly while maintaining the test performance of the remaining two as per the base case, and simultaneously for all components. The base case average net monetary benefits were $1·49 and -$0 ·38 in Cambodia and Bangladesh, respectively. CRP, C-reactive protein; LF-RDT, lateral flow rapid diagnostic test. Costs were adjusted for inflation and expressed in 2022 US dollars. *The average net monetary benefit was calculated based on a multiplex LF-RDT price of $5 per unit.

| Multiplex LF-RDT component | Sensitivity, % | Specificity, % | Cambodia |  | Bangladesh |  |
| --- | --- | --- | --- | --- | --- | --- |
| | | | Average net monetary benefit*, US\$ | Threshold cost-effective price, US\$ | Average net monetary benefit*, US\$ | Threshold cost-effective price, US\$ |
| Enteric fever diagnostic test | 90 | 60 | -1.40 | 3.60 | -2.23 | 2.77 |
|  | 80 | 70 | -0.70 | 4.30 | -2.11 | 2.89 |
|  | 70 | 80 | 0.01 | 5.01 | -1.99 | 3.01 |
|  | 60 | 90 | 0.72 | 5.72 | -1.87 | 3.13 |
| Dengue diagnostic test | 90 | 60 | -0.85 | 4.15 | -1.34 | 3.66 |
|  | 80 | 70 | -0.07 | 4.93 | -0.97 | 4.03 |
|  | 70 | 80 | 0.71 | 5.71 | -0.60 | 4.40 |
|  | 60 | 90 | 1.49 | 6.49 | -0.24 | 4.76 |
| CRP test | 90 | 60 | 0.48 | 5.48 | -1.18 | 3.82 |
|  | 80 | 70 | 0.93 | 5.93 | -0.82 | 4.18 |
|  | 70 | 80 | 1.39 | 6.39 | -0.45 | 4.55 |
|  | 60 | 90 | 1.84 | 6.84 | -0.09 | 4.91 |
| All components | 90 | 60 | -4.09 | 0.91 | -3.49 | 1.51 |
|  | 80 | 70 | -2.54 | 2.46 | -2.93 | 2.07 |
|  | 70 | 80 | -0.83 | 4.17 | -2.25 | 2.75 |
|  | 60 | 90 | 1.04 | 6.04 | -1.44 | 3.56 |

Using the multiplex LF-RDT only on patients with fever with no localizing symptoms improved cost-effectiveness greatly, with the intervention remaining dominant in Cambodia and below the lower willingness-to-pay threshold in Bangladesh with an ICER of $145 per DALY averted. Unsurprisingly, increasing the mortality benefit of appropriately diagnosing and treating enteric fever from 0.25% to 0.5% also increased cost-effectiveness, with the cost per DALY averted in Bangladesh reduced to $267, lower than the conservative willingness-to-pay threshold of $388. This cost was multiplied nearly ten-fold to $2,417 per DALY averted were there no mortality benefit, which considerably exceeds the lower threshold but remains below the upper per capita GDP-based threshold of $2,687. In Cambodia, however, multiplex LF-RDT-guided clinical management remained dominant regardless of whether it had an effect on enteric fever mortality, reflecting the findings of the deterministic sensitivity analysis. The dominance of the intervention in Cambodia was also seen when the cost of antimicrobial resistance was excluded from the model, whereas this raised the ICER in Bangladesh from $482 to $579 per DALY averted, which is still clearly under the upper willingness-to-pay threshold.

## DISCUSSION

The main finding from this study is that a putative multiplex LF-RDT using capillary blood as the test substrate with the ability to diagnose enteric fever and dengue, and measure CRP to distinguish between other viral and non-viral causes of acute fever would be dominant in Cambodia. In Bangladesh, the multiplex LF-RDT would be considered cost-effective with respect to the widely used per capita GDP willingness-to-pay threshold, but not with respect to the more conservative opportunity cost-based threshold. This is borne out by the results of the probabilistic sensitivity analyses. Despite the number of DALYs averted through the use of the multiplex LF-RDT in Bangladesh being more than triple that in Cambodia, as reflected by higher disease incidences in the former, cost-savings were fewer due to the lower societal cost of antimicrobial resistance and lower cost of healthcare provision in both hospital and primary care settings. As a result, the average net monetary benefit per patient tested of more judicious antibiotic prescribing, avoiding hospital admission in those testing negative for dengue, and more accurate diagnosis and management of enteric fever were unable to exceed the average total cost, 55% of which comprised the $5 price of a multiplex LF-RDT. However, it should be noted that the estimates of the cost-effectiveness of the multiplex LF-RDT produced in this analysis were conservative. For one, only a modest health benefit from improved diagnosis was incorporated in the model i.e., reduced hospitalisations and mortality from enteric fever, with no other mortality or morbidity benefit included for improved diagnosis and treatment of dengue and other bacterial or viral infections. Given the low levels of incremental costs, even further small increases in the estimated incremental DALYs averted would increase the cost-effectiveness of the multiplex LF-RDT substantially, as can be seen in Bangladesh where a reduction of enteric fever mortality in the intervention arm of the model from the base case figure of 0·75% to 0·5% drove the cost per DALY averted considerably under the conservative opportunity-cost based willingness-to-pay threshold.

A key strength of this study is the use of data from the largest study on the epidemiology of acute febrile illness in rural South and Southeast Asia,^20^ combined with other country- or region-specific data, to parameterize the model. Furthermore, the model factors in the societal cost of antimicrobial resistance, as it is crucial for policymakers that this cost is accounted for when evaluating the cost-effectiveness of interventions which have the potential to improve antimicrobial stewardship, notwithstanding the methodological challenges. This is especially true for South and Southeast Asian countries where antimicrobial resistance is an urgent public health issue,^54^ and where simple, low-cost interventions such as this multiplex LF-RDT may be considered for integration into national or sub-national strategic plans targeting antimicrobial resistance. We have shown its potential to shift the burden of inappropriate prescribing from the high societal-cost antibiotic ciprofloxacin to the relatively lower societal-cost amoxicillin, despite only modest reductions in overall antibiotic prescription rates.

Nonetheless, this study has several limitations. First, the data on which costs of healthcare provision in Bangladesh are based are not contemporary and, thus, may be much higher than estimated, especially given advances at the secondary care level. Were this to be the case, then the multiplex LF-RDT would tend towards being cost-effective in Bangladesh, since hospitalisation costs comprise the bulk of the total costs there. Second, due to an absence of robust data it was assumed that augmenting clinical assessment with the multiplex LF-RDT resulted in only a very small absolute reduction in mortality from enteric fever and no other mortality or morbidity difference between the two arms of the model, but it is conceivable that earlier diagnosis and better directed therapy for other bacterial infections would reduce complications including mortality. In both settings, this would increase the number of DALYs averted, again predisposing the intervention towards cost-effectiveness, an effect which would be of more importance in Bangladesh. A more complex model which extends the one-year time horizon and captures transitions between health states may give more detailed results than our conservative estimates. Third, no official willingness-to-pay thresholds set by policymakers against which the cost-effectiveness of the multiplex LF-RDT could be benchmarked were available; additionally, it should be recognized that there is an element of subjectivity and different approaches to threshold-setting. Lastly, the model was developed based on 12 months’ worth of epidemiological data. Thus, while inter-year seasonal variation in acute NMFI incidence was accounted for, it was not possible for inter-year variation to be similarly considered.

In previous work, we have established the urgency of the need for such a multiplex LF-RDT in rural South and Southeast Asia.^10^ However, no evaluation of its potential cost-effectiveness has been conducted, much less at a country level. The principal contribution of our findings, therefore, is that they provide guidance for researchers and industry partners working on diagnostics for use in LMICs in terms of performance characteristic and pricing targets, as well as providing policymakers with a baseline economic viewpoint on which to make decisions concerning such tests in future. For example, the price of the multiplex LF-RDT could be set higher in high-income countries, should it be marketed there, in order to lower the price in LMICs, thus ensuring cost-effectiveness in as many countries as possible.

Our results bolster the limited evidence provided by other cost-effectiveness analyses in South and Southeast Asian primary care settings of LF-RDTs which singularly test for enteric fever, dengue, and CRP (or other host biomarker associated with bacterial infection). In two modelling studies set in rural Laos, the pathogen-agnostic nature of CRP was likely to be highly cost-effective despite heterogeneity in the causes of NMFI. This was true when an antibiotic prescribing threshold of 40 mg/L, similar to that used in this study, was applied, and also when a more liberal threshold of 20 mg/L was used.^25,27^ In one of these studies, a dengue LF-RDT with both sensitivity and specificity of 95% was dominant.^25^ Another modelling study of a commercially-available typhoid IgM LF-RDT in Cambodian children with suspected enteric fever did not find it to be cost-effective, but used an intermediate outcome measure (additional treatment success), rather than DALY averted. Moreover, the sensitivity of this test was only 59%, as opposed to 85% in this study.^26^ There are even fewer studies pertaining to South Asia, but a modelling study of CRP-guided antibiotic prescription in Afghan primary care facilities showed it to be highly cost-effective with an incremental cost of $14 per additional correctly treated case, even with a very low cut-off of 10 mg/L.^28^ Notably, the costs of the tests used in these studies were all <$5. Considering the results of our study in the light of these others, the multiplex LF-RDT would likely be cost-effective in at least several other South and Southeast Asian countries, especially given the high prevalences of enteric fever and dengue in the region. For instance, a shift from diagnosing dengue clinically, which priorities sensitivity, to diagnosis via the multiplex LF-RDT, which prioritises specificity, would prevent over-referral to hospital which would reduce secondary care costs and ensure capacity during periods of high demand, such as dengue outbreaks.

Furthermore, in view of the likely constraint on sensitivity imposed by small substrate volumes, we have determined through the first scenario analysis the components of the multiplex LF-RDT which could preferentially have their sensitivities reduced while optimising cost-effectiveness i.e., dengue and CRP tests. We have also identified several key areas for future study in order to improve the informativeness of the analysis for policymaking, one of which is contemporaneous quantification of healthcare provision costs in Bangladesh. In addition, since it is unlikely that this multiplex LF-RDT will be developed and rolled out at scale in the very near future, the results of the second scenario analysis indicate that upskilling rural primary health workers in Cambodia and Bangladesh in formulating syndromic diagnoses in the interim will be beneficial. This is because not only will the total spend on test kits be reduced, but cost-effectiveness will also be increased if they were only used for patients with fever but no localizing symptoms; however, primary health workers first need to be trained to identify this subset of patients. This finding may also be applicable to other countries in South and Southeast Asia with similar acute febrile illness epidemiology and health system developmental status. Context-specific evaluations of the key drivers of cost-effectiveness identified in this study will further aid assessment of the generalizability of the cost-effectiveness of this multiplex LF-RDT to other countries, as will consideration of other contextual factors such as healthcare worker adherence to test-guided clinical management when the multiplex LF-RDT has been developed.

## CONCLUSION

Our conservative model-based cost-effectiveness analysis of a multiplex LF-RDT to aid diagnosis and management of patients with NMFI has shown the potential for this intervention to be cost-effective under the right context-specific conditions. This should provide much-needed impetus from an economic perspective for the development of this multiplex LF-RDT, encourage further studies to determine the optimal configuration of conditions in South and Southeast Asian countries where it may potentially be used, and, where possible, guide adjustment of these e.g., country-specific unit prices to ensure cost-effectiveness.

## Author contributions

RC – conceptualization, methodology, investigation, formal analysis, visualization, writing – original draft; CP – methodology, investigation, formal analysis, visualization, writing – review and editing; WP – supervision, writing – review and editing; NPJD – supervision, funding acquisition, writing – review and editing; YL – methodology, supervision, funding acquisition, writing – review and editing

## ACKNOWLEDGEMENTS

We thank Action for Health and Development (AHEAD) and the Battambang Provincial Health Department for providing data on antibiotic costs in Cambodia.

## DECLARATIONS

### Ethics approval

Ethical approval was obtained from the University of Oxford Tropical Research Ethics Committee (OxTREC/543-20), the Cambodian National Ethics Committee for Health Research (125/NECHR), and the Bangladesh Medical Research Council Ethics Committee (BMRC/NREC/2019-2022/133).

### Competing interests

The authors declare no competing interests.

### Data availability

All data relevant to the study are included in the article or uploaded as supplementary information.

### Funding

This research was funded in whole, or in part, by the Wellcome Trust [215604/Z/19/Z]. RC was also funded by the UK Government through a Commonwealth Scholarship, the Royal Australasian College of Physicians through the Bushell Travelling Fellowship in Medicine or the Allied Sciences, and the Rotary Foundation through a Global Grant Scholarship.

The funders had no role in study design, data collection, data analysis, data interpretation, or writing of the manuscript. All authors had full access to the data and had final responsibility for the decision to submit the manuscript for publication. For the purpose of open access, the authors have applied a CC BY public copyright licence to any Author Accepted Manuscript version arising from this submission.

## Notes

### Competing Interest Statement

The authors have declared no competing interest.

### Author Declarations

The University of Oxford Tropical Research Ethics Committee (OxTREC/543-20), the Cambodian National Ethics Committee for Health Research (125/NECHR), and the Bangladesh Medical Research Council Ethics Committee (BMRC/NREC/2019-2022/133) gave ethical approval for this work.

