## Supporting Information S1 for "Model-based cost-effectiveness analysis of a multiplex lateral flow rapid diagnostic test for acute non-malarial febrile illness in rural South and Southeast Asian primary care"

### RECOMMENDED INTERPRETATION AND MANAGEMENT FOR POSITIVE TESTS

| Positive multiplex LF-RDT component | Recommended interpretation | Recommended management |
|---|---|---|
| Dengue | Patient has dengue | Referral to hospital for admission |
| Enteric fever | Patient has enteric fever | Ciprofloxacin |
| CRP (>40 mg/L) | Patient has a bacterial infection (not enteric fever if enteric fever test is negative, otherwise enteric fever if enteric fever test also positive) | Amoxicillin unless enteric fever test also positive, in which case ciprofloxacin |
| None | Patient has a viral infection | No antibiotics |

### CAMBODIA

#### Model Settings

Reset all model inputs

Introduction

Settings

Results

SA Settings

DSA Results

PSA Results

**Model settings**

| | | |
|---|---|---|
| Time horizon | Lifetime | |
| DSA/PSA default variation | 20.00% | |
| Number of PSA iterations | 1,000 | |
| Willingness-to-pay threshold<br>Note: Adjust if changing country | $357 | *Woods et al. Inflated to 2022 values - Bangladesh: 51-723 (avg 388). Cambodia: 56-658 (avg 357). Original values - Bangladesh: 30-427 (avg 229). Cambodia: 44-518 (avg 281). 2013 data.* |
| Perspective | Societal | |
| Country | Cambodia | |

**Population characteristics**

| | Value | Source |
|---|---|---|
| Average age | 28 | SEACTN data (21/03/2022-21/03/2023 - Bangladesh: 28.34 ;Cambodia: 34.21) |
| Proportion of adults | 79.05% | SEACTN data (21/03/2022-21/03/2023) |
| Proportion of children 5-15 years | 17.14% | SEACTN data (21/03/2022-21/03/2023) |
| Proportion of children <5 years | 3.81% | SEACTN data (21/03/2022-21/03/2023) |

**Scenario analysis inputs (in place of the above)**

Fever of unknown source

| | | |
|---|---|---|
| Proportion of adults | 82.84% | SEACTN data (21/03/2022-21/03/2023) |
| Proportion of children 5-15 years | 13.64% | SEACTN data (21/03/2022-21/03/2023) |
| Proportion of children <5 years | 3.52% | SEACTN data (21/03/2022-21/03/2023) |

### Decision Tree

Introduction
Settings
Results
SA Settings
DSA Results
PSA Results

**Decision tree calculations**

**Intervention:** Clinical assessment only

| Branch | True disease | Diagnosis | Outcome | Probability | Diagnostic costs | Hospitalisation costs | Treatment costs | Societal costs | **Total costs** | YLD | YLL | **DALYs** |
|---|---|---|---|---|---|---|---|---|---|---|---|---|
| 1 | Enteric fever | Enteric fever | Survive | 1.27% | $0.00 | $0.23 | $0.02 | $0.06 | **$0.31** | 0.0094 | *0.0000* | **0.0094** |
| 2 | Enteric fever | Enteric fever | Die | 0.01% | $0.00 | $0.00 | $0.00 | $0.00 | **$0.00** | 0.0001 | 0.0038 | **0.0039** |
| 3 | Enteric fever | Other | Survive | 1.26% | $0.00 | $0.37 | $0.03 | $0.02 | **$0.42** | 0.0098 | *0.0000* | **0.0098** |
| 4 | Enteric fever | Other | Die | 0.01% | $0.00 | $0.00 | $0.00 | $0.00 | **$0.00** | 0.0001 | 0.0051 | **0.0052** |
| 5 | Dengue | Dengue | Survive - observed in hospital | 11.56% | $0.00 | $1.20 | *$0.00* | *$0.00* | **$1.20** | *0.0000* | *0.0000* | **0.0000** |
| 6 | Dengue | Other | Survive - not observed in hospital | 0.87% | $0.00 | $0.00 | *$0.00* | *$0.00* | **$0.00** | *0.0000* | *0.0000* | **0.0000** |
| 7 | Other viral | Enteric fever | Survive | 17.28% | $0.00 | $1.19 | $0.29 | $0.80 | **$2.28** | *0.0000* | *0.0000* | **0.0000** |
| 8 | Other viral | Dengue | Survive | 47.69% | $0.00 | $4.94 | *$0.00* | *$0.00* | **$4.94** | *0.0000* | *0.0000* | **0.0000** |
| 9 | Other viral | Other bacterial | Survive - antibiotic prescribed | 1.79% | $0.00 | $0.00 | $0.06 | $0.04 | **$0.10** | *0.0000* | *0.0000* | **0.0000** |
| 10 | Other viral | Other viral | Survive - no antibiotic prescribed | 2.36% | $0.00 | $0.00 | *$0.00* | *$0.00* | **$0.00** | *0.0000* | *0.0000* | **0.0000** |
| 11 | Other bacterial cause | Other bacterial | Survive - antibiotic prescribed | 11.16% | $0.00 | $0.00 | $0.37 | $0.28 | **$0.64** | *0.0000* | *0.0000* | **0.0000** |
| 12 | Other bacterial cause | Other viral | Survive - no antibiotic prescribed | 4.74% | $0.00 | $0.00 | *$0.00* | *$0.00* | **$0.00** | *0.0000* | *0.0000* | **0.0000** |
| | | | | **100.00%** | **$0.00** | **$7.93** | **$0.76** | **$1.21** | **$9.90** | **0.0194** | **0.0089** | **0.0282** |

| | |
|---|---|
| Percentage receiving antibiotics | 31.51% |
| Inappropriate antibiotics | 20.02% |

**Intervention:** Novel multiplex + clinical assessment

| Branch | True disease | Diagnosis | Outcome | Probability | Diagnostic costs | Hospitalisation costs | Treatment costs | Societal costs | **Total costs** | YLD | YLL | **DALYs** |
|---|---|---|---|---|---|---|---|---|---|---|---|---|
| 1 | Enteric fever | Enteric fever | Survive | 2.15% | $0.11 | $0.39 | $0.04 | $0.10 | **$0.63** | 0.0160 | *0.0000* | **0.0160** |
| 2 | Enteric fever | Enteric fever | Die | 0.02% | $0.00 | $0.00 | $0.00 | $0.00 | **$0.00** | 0.0001 | 0.0065 | **0.0066** |
| 3 | Enteric fever | Other | Survive | 0.38% | $0.02 | $0.11 | $0.01 | $0.01 | **$0.15** | 0.0029 | *0.0000* | **0.0029** |
| 4 | Enteric fever | Other | Die | 0.00% | $0.00 | $0.00 | $0.00 | $0.00 | **$0.00** | 0.0000 | 0.0015 | **0.0016** |
| 5 | Dengue | Dengue | Survive - observed in hospital | 10.56% | $0.53 | $1.09 | *$0.00* | *$0.00* | **$1.62** | *0.0000* | *0.0000* | **0.0000** |
| 6 | Dengue | Other | Survive - not observed in hospital | 1.86% | $0.09 | $0.00 | *$0.00* | *$0.00* | **$0.09** | *0.0000* | *0.0000* | **0.0000** |
| 7 | Other viral | Enteric fever | Survive | 3.46% | $0.17 | $0.24 | $0.06 | $0.16 | **$0.63** | *0.0000* | *0.0000* | **0.0000** |
| 8 | Other viral | Dengue | Survive | 3.46% | $0.17 | $0.36 | *$0.00* | *$0.00* | **$0.53** | *0.0000* | *0.0000* | **0.0000** |
| 9 | Other viral | Other bacterial | Survive - antibiotic prescribed | 9.95% | $0.50 | $0.00 | $0.33 | $0.25 | **$1.07** | *0.0000* | *0.0000* | **0.0000** |
| 10 | Other viral | Other viral | Survive - no antibiotic prescribed | 52.25% | $2.61 | $0.00 | *$0.00* | *$0.00* | **$2.61** | *0.0000* | *0.0000* | **0.0000** |
| 11 | Other bacterial cause | Other bacterial | Survive - antibiotic prescribed | 11.77% | $0.59 | $0.00 | $0.39 | $0.29 | **$1.27** | *0.0000* | *0.0000* | **0.0000** |
| 12 | Other bacterial cause | Other viral | Survive - no antibiotic prescribed | 4.13% | $0.21 | $0.00 | *$0.00* | *$0.00* | **$0.21** | *0.0000* | *0.0000* | **0.0000** |
| | | | | **100.00%** | **$5.00** | **$2.20** | **$0.81** | **$0.81** | **$8.82** | **0.0191** | **0.0080** | **0.0271** |

| | |
|---|---|
| Percentage receiving antibiotics | 27.34% |
| Inappropriate antibiotics | 13.69% |

### Model Results

Introduction
Settings
Results
SA Settings
DSA Results
PSA Results

**Cost-Effectiveness Results**

| | Diagnostic costs | Hospitalisation costs | Treatment costs | Societal costs | **Total Costs** | YLD | YLL | **DALYs** | % of cases correctly diagnosed | % of patients prescribed antibiotics |
|---|---|---|---|---|---|---|---|---|---|---|
| Clinical assessment | $0.00 | $7.93 | $0.76 | $1.21 | **$9.90** | 0.0194 | 0.0089 | **0.0282** | 32.88% | 31.51% |
| Multiplex LF-RDT | $5.00 | $2.20 | $0.81 | $0.81 | **$8.82** | 0.0191 | 0.0080 | **0.0271** | 90.84% | 27.34% |

**Incremental Results**

| | Diagnostic costs | Hospitalisation costs | Treatment costs | Societal costs | **Total Costs** | YLD | YLL | **DALYs averted** | % of cases correctly diagnosed | % of patients prescribed antibiotics | **ICER ($/DALY averted)** |
|---|---|---|---|---|---|---|---|---|---|---|---|
| Multiplex LF-RDT vs. Clinical assessment | $5.00 | -$5.74 | $0.05 | -$0.40 | **-$1.08** | 0.0003 | 0.0009 | **0.0012** | 57.96% | -4.16% | -$940.95 |

l

### Scenario Analyses

Run Scenario

Introduction

Settings

Results

SA Settings

DSA Results

PSA Results

**Sensitivity and Specificity Scenario Analyses**

| Dengue multiplex | | | |
|---|---|---|---|
| Sensitivity | Specificity | NMB | Threshold cost-effective price |
| 0.9 | 0.6 | $1.42 | $6.42 |
| 0.8 | 0.7 | $2.05 | $7.05 |
| 0.7 | 0.8 | $2.69 | $7.69 |
| 0.6 | 0.9 | $3.33 | $8.33 |

NMB $1.49

| Enteric fever multiplex | | | |
|---|---|---|---|
| Sensitivity | Specificity | NMB | Threshold cost-effective price |
| 0.9 | 0.6 | -$5.08 | -$0.08 |
| 0.8 | 0.7 | -$2.71 | $2.29 |
| 0.7 | 0.8 | -$0.34 | $4.66 |
| 0.6 | 0.9 | $2.03 | $7.03 |

| CRP multiplex | | | |
|---|---|---|---|
| Sensitivity | Specificity | NMB | Threshold cost-effective price |
| 0.9 | 0.6 | -$0.04 | $4.96 |
| 0.8 | 0.7 | $1.44 | $6.44 |
| 0.7 | 0.8 | $2.91 | $7.91 |
| 0.6 | 0.9 | $4.39 | $9.39 |

| All 3 tests | | | |
|---|---|---|---|
| Sensitivity | Specificity | NMB | Threshold cost-effective price |
| 0.9 | 0.6 | -$8.03 | -$3.03 |
| 0.8 | 0.7 | -$4.83 | $0.17 |
| 0.7 | 0.8 | -$1.09 | $3.91 |
| 0.6 | 0.9 | $3.17 | $8.17 |

### BANGLADESH

#### Model Settings

Reset all model inputs

Introduction

Settings

Results

SA Settings

DSA Results

PSA Results

**Model settings**

| | | |
|---|---|---|
| Time horizon | Lifetime | |
| DSA/PSA default variation | 20.00% | |
| Number of PSA iterations | 1,000 | |
| Willingness-to-pay threshold<br>Note: Adjust if changing country | $388 | *Woods et al. Inflated to 2022 values - Bangladesh: 51-723 (avg 388). Cambodia: 56-658 (avg 357). Original values - Bangladesh: 30-427 (avg 229). Cambodia: 44-518 (avg 281). 2013 data.* |
| Perspective | Societal | |
| Country | Bangladesh | |

**Population characteristics**

| | Value | Source |
|---|---|---|
| Average age | 28 | SEACTN data (21/03/2022-21/03/2023 - Bangladesh: 28.34 ;Cambodia: 34.21) |
| Proportion of adults | 79.05% | SEACTN data (21/03/2022-21/03/2023) |
| Proportion of children 5-15 years | 17.14% | SEACTN data (21/03/2022-21/03/2023) |
| Proportion of children <5 years | 3.81% | SEACTN data (21/03/2022-21/03/2023) |

**Scenario analysis inputs (in place of the above)**

Fever of unknown source

| | | |
|---|---|---|
| Proportion of adults | 82.84% | SEACTN data (21/03/2022-21/03/2023) |
| Proportion of children 5-15 years | 13.64% | SEACTN data (21/03/2022-21/03/2023) |
| Proportion of children <5 years | 3.52% | SEACTN data (21/03/2022-21/03/2023) |

### Decision Tree

Introduction
Settings
Results
SA Settings
DSA Results
PSA Results

**Decision tree calculations**

**Intervention:** Clinical assessment only

| Branch | True disease | Diagnosis | Outcome | Probability | Diagnostic costs | Hospitalisation costs | Treatment costs | Societal costs | **Total costs** | YLD | YLL | **DALYs** |
|---|---|---|---|---|---|---|---|---|---|---|---|---|
| 1 | Enteric fever | Enteric fever | Survive | 1.27% | $0.00 | $0.23 | $0.02 | $0.06 | **$0.31** | 0.0094 | *0.0000* | **0.0094** |
| 2 | Enteric fever | Enteric fever | Die | 0.01% | $0.00 | $0.00 | $0.00 | $0.00 | **$0.00** | 0.0001 | 0.0038 | **0.0039** |
| 3 | Enteric fever | Other | Survive | 1.26% | $0.00 | $0.37 | $0.03 | $0.02 | **$0.42** | 0.0098 | *0.0000* | **0.0098** |
| 4 | Enteric fever | Other | Die | 0.01% | $0.00 | $0.00 | $0.00 | $0.00 | **$0.00** | 0.0001 | 0.0051 | **0.0052** |
| 5 | Dengue | Dengue | Survive - observed in hospital | 11.56% | $0.00 | $1.20 | *$0.00* | *$0.00* | **$1.20** | *0.0000* | *0.0000* | **0.0000** |
| 6 | Dengue | Other | Survive - not observed in hospital | 0.87% | $0.00 | $0.00 | *$0.00* | *$0.00* | **$0.00** | *0.0000* | *0.0000* | **0.0000** |
| 7 | Other viral | Enteric fever | Survive | 17.28% | $0.00 | $1.19 | $0.29 | $0.80 | **$2.28** | *0.0000* | *0.0000* | **0.0000** |
| 8 | Other viral | Dengue | Survive | 47.69% | $0.00 | $4.94 | *$0.00* | *$0.00* | **$4.94** | *0.0000* | *0.0000* | **0.0000** |
| 9 | Other viral | Other bacterial | Survive - antibiotic prescribed | 1.79% | $0.00 | $0.00 | $0.06 | $0.04 | **$0.10** | *0.0000* | *0.0000* | **0.0000** |
| 10 | Other viral | Other viral | Survive - no antibiotic prescribed | 2.36% | $0.00 | $0.00 | *$0.00* | *$0.00* | **$0.00** | *0.0000* | *0.0000* | **0.0000** |
| 11 | Other bacterial cause | Other bacterial | Survive - antibiotic prescribed | 11.16% | $0.00 | $0.00 | $0.37 | $0.28 | **$0.64** | *0.0000* | *0.0000* | **0.0000** |
| 12 | Other bacterial cause | Other viral | Survive - no antibiotic prescribed | 4.74% | $0.00 | $0.00 | *$0.00* | *$0.00* | **$0.00** | *0.0000* | *0.0000* | **0.0000** |
| | | | | **100.00%** | **$0.00** | **$7.93** | **$0.76** | **$1.21** | **$9.90** | **0.0194** | **0.0089** | **0.0282** |

Percentage receiving antibiotics 31.51%
Inappropriate antibiotics 20.02%

**Intervention:** Novel multiplex + clinical assessment

| Branch | True disease | Diagnosis | Outcome | Probability | Diagnostic costs | Hospitalisation costs | Treatment costs | Societal costs | **Total costs** | YLD | YLL | **DALYs** |
|---|---|---|---|---|---|---|---|---|---|---|---|---|
| 1 | Enteric fever | Enteric fever | Survive | 2.15% | $0.11 | $0.39 | $0.04 | $0.10 | **$0.63** | 0.0160 | *0.0000* | **0.0160** |
| 2 | Enteric fever | Enteric fever | Die | 0.02% | $0.00 | $0.00 | $0.00 | $0.00 | **$0.00** | 0.0001 | 0.0065 | **0.0066** |
| 3 | Enteric fever | Other | Survive | 0.38% | $0.02 | $0.11 | $0.01 | $0.01 | **$0.15** | 0.0029 | *0.0000* | **0.0029** |
| 4 | Enteric fever | Other | Die | 0.00% | $0.00 | $0.00 | $0.00 | $0.00 | **$0.00** | 0.0000 | 0.0015 | **0.0016** |
| 5 | Dengue | Dengue | Survive - observed in hospital | 10.56% | $0.53 | $1.09 | *$0.00* | *$0.00* | **$1.62** | *0.0000* | *0.0000* | **0.0000** |
| 6 | Dengue | Other | Survive - not observed in hospital | 1.86% | $0.09 | $0.00 | *$0.00* | *$0.00* | **$0.09** | *0.0000* | *0.0000* | **0.0000** |
| 7 | Other viral | Enteric fever | Survive | 3.46% | $0.17 | $0.24 | $0.06 | $0.16 | **$0.63** | *0.0000* | *0.0000* | **0.0000** |
| 8 | Other viral | Dengue | Survive | 3.46% | $0.17 | $0.36 | *$0.00* | *$0.00* | **$0.53** | *0.0000* | *0.0000* | **0.0000** |
| 9 | Other viral | Other bacterial | Survive - antibiotic prescribed | 9.95% | $0.50 | $0.00 | $0.33 | $0.25 | **$1.07** | *0.0000* | *0.0000* | **0.0000** |
| 10 | Other viral | Other viral | Survive - no antibiotic prescribed | 52.25% | $2.61 | $0.00 | *$0.00* | *$0.00* | **$2.61** | *0.0000* | *0.0000* | **0.0000** |
| 11 | Other bacterial cause | Other bacterial | Survive - antibiotic prescribed | 11.77% | $0.59 | $0.00 | $0.39 | $0.29 | **$1.27** | *0.0000* | *0.0000* | **0.0000** |
| 12 | Other bacterial cause | Other viral | Survive - no antibiotic prescribed | 4.13% | $0.21 | $0.00 | *$0.00* | *$0.00* | **$0.21** | *0.0000* | *0.0000* | **0.0000** |
| | | | | **100.00%** | **$5.00** | **$2.20** | **$0.81** | **$0.81** | **$8.82** | **0.0191** | **0.0080** | **0.0271** |

Percentage receiving antibiotics 27.34%
Inappropriate antibiotics 13.69%

### Model Results

Introduction

Settings

Results

SA Settings

DSA Results

PSA Results

**Cost-Effectiveness Results**

| | Diagnostic costs | Hospitalisation costs | Treatment costs | Societal costs | **Total Costs** | YLD | YLL | **DALYs** | % of cases correctly diagnosed | % of patients prescribed antibiotics |
|---|---|---|---|---|---|---|---|---|---|---|
| Clinical assessment | $0.00 | $7.93 | $0.76 | $1.21 | **$9.90** | 0.0194 | 0.0089 | **0.0282** | 32.88% | 31.51% |
| Multiplex LF-RDT | $5.00 | $2.20 | $0.81 | $0.81 | **$8.82** | 0.0191 | 0.0080 | **0.0271** | 90.84% | 27.34% |

**Incremental Results**

| | Diagnostic costs | Hospitalisation costs | Treatment costs | Societal costs | **Total Costs** | YLD | YLL | **DALYs averted** | % of cases correctly diagnosed | % of patients prescribed antibiotics | **ICER ($/DALY averted)** |
|---|---|---|---|---|---|---|---|---|---|---|---|
| Multiplex LF-RDT vs. Clinical assessment | $5.00 | -$5.74 | $0.05 | -$0.40 | **-$1.08** | 0.0003 | 0.0009 | **0.0012** | 57.96% | -4.16% | -$940.95 |

l

### Scenario Analyses

Run Scenario

Introduction

Settings

Results

SA Settings

DSA Results

PSA Results

**Sensitivity and Specificity Scenario Analyses**

NMB $1.49

| Dengue multiplex | | | |
|---|---|---|---|
| Sensitivity | Specificity | NMB | Threshold cost-effective price |
| 0.9 | 0.6 | $1.42 | $6.42 |
| 0.8 | 0.7 | $2.05 | $7.05 |
| 0.7 | 0.8 | $2.69 | $7.69 |
| 0.6 | 0.9 | $3.33 | $8.33 |

| Enteric fever multiplex | | | |
|---|---|---|---|
| Sensitivity | Specificity | NMB | Threshold cost-effective price |
| 0.9 | 0.6 | -$5.08 | -$0.08 |
| 0.8 | 0.7 | -$2.71 | $2.29 |
| 0.7 | 0.8 | -$0.34 | $4.66 |
| 0.6 | 0.9 | $2.03 | $7.03 |

| CRP multiplex | | | |
|---|---|---|---|
| Sensitivity | Specificity | NMB | Threshold cost-effective price |
| 0.9 | 0.6 | -$0.04 | $4.96 |
| 0.8 | 0.7 | $1.44 | $6.44 |
| 0.7 | 0.8 | $2.91 | $7.91 |
| 0.6 | 0.9 | $4.39 | $9.39 |

| All 3 tests | | | |
|---|---|---|---|
| Sensitivity | Specificity | NMB | Threshold cost-effective price |
| 0.9 | 0.6 | -$8.03 | -$3.03 |
| 0.8 | 0.7 | -$4.83 | $0.17 |
| 0.7 | 0.8 | -$1.09 | $3.91 |
| 0.6 | 0.9 | $3.17 | $8.17 |
